# Specific long-term changes in anti-SARS-CoV-2 IgG modifications and antibody functions in mRNA, adenovector, and protein subunit vaccines

**DOI:** 10.1101/2023.06.16.23291455

**Authors:** Sebastian Reinig, Chin Kuo, Chia-Chun Wu, Sheng-Yu Huang, Jau-Song Yu, Shin-Ru Shih

## Abstract

Various vaccine platforms were developed and deployed against the COVID-19 disease. The Fc-mediated functions of IgG antibodies are essential in the adaptive immune response elicited by vaccines. However, the long-term changes of protein subunit vaccines and their combinations with mRNA vaccines are unknown. A total of 272 serum and plasma samples were collected from individuals who received first to third doses of the protein subunit Medigen, the mRNA (BNT), or the adenovector AstraZeneca vaccines. The IgG subclass level was measured using enzyme-linked immunosorbent assay, and Fc-N glycosylation was measured using LC-MS/MS. Antibody-dependent phagocytosis (ADCP) and complement deposition (ADCD) of anti-spike (S) IgG antibodies were measured. IgG1 and 3 reached the highest anti-S IgG subclass level. IgG1, 2, and 4 subclass levels significantly increased in mRNA- and Medigen-vaccinated individuals. Fc-glycosylation was stable, except in female BNT vaccinees, who showed increased bisection and decreased galactosylation. Female BNT vaccinees had a higher anti-S IgG titer than that of males. ADCP declined in all groups. ADCD increased in Medigen-vaccinated individuals after the third dose. Each vaccine produced specific long-term changes in Fc structure and function. This finding is critical when selecting a vaccine platform or combination to achieve the desired immune response.

## Introduction

The novel severe acute respiratory syndrome coronavirus 2 (SARS-CoV-2) causes the coronavirus disease 2019 (COVID-19) pandemic and has sparked the rapid development of various vaccine platforms^1^. Antigenic antibody generation against the virus is a primary component of the immune response and memory evoked by vaccination. The IgG antibody is one of the primary humoral responses of the vaccines^2,3^ evoked by COVID-19 vaccines. Many different vaccine platforms (except inactivated vaccines), including genetic vaccines like mRNA and adenovector vaccines, and classical vaccine platforms like protein subunit vaccines, were developed to target the spike (S)-glycoprotein as a single antigen. The development of these vaccine platforms enables comparisons of the effect of the vaccine platform on the immune response.

Most studies focused on neutralizing antibodies that block viral entry. While neutralizing IgG antibodies correlates with protection against COVID-19 on a population level, neutralizing IgG antibodies are insufficient to explain the protection elicit by the vaccines since many vaccines can still lead to infection and symptoms despite high neutralizing antibody titers^3^. IgG can also bind to the Fc receptor and activate the innate immune effector via its Fc domain, including antibody-dependent phagocytosis (ADCP), cellular cytotoxicity (ADCC), and complement deposition (ADCD)^4^. Fc-mediated functions are potentially the primary mechanisms by which IgG antibodies are activated against the SARS-CoV-2 virus. The affinity of IgG for Fc-mediated functions is determined by the isotype (subclass) and the conserved n-297 glycosylation site^5,6^. Several studies have investigated the long-term changes in the IgG profile of mRNA vaccines, showing an increased contribution of the IgG4 subclass of anti-S IgG^7–9^ associated with weaker Fc-mediated functions^7^ (7). In Taiwan, a domestically developed protein subunit vaccine against COVID-19 was developed and administered (Medigen MVC-COV1901)^10,11^. However, the long-term immune changes elicited by protein subunit vaccines and the effects of combining them with mRNA vaccines are still unclear. Therefore, in this study, we measured and compared the long-term changes in anti-S-IgG subclasses, Fc-glycosylation, and ADCP and ADCD function for 1–3 vaccine doses for over one year after initial immunization of the protein subunit, adenovectors, and mRNA vaccines. This study is the first to investigate the antigenic IgG profile and antibody functions of the Medigen protein subunit vaccine in humans.

## Subjects and Methods

### Sample collection

Serum and plasma samples were obtained from COVID-19-vaccinated individuals at Chang Gung University and Chang Gung Memorial Hospital (**Table I**). Samples were collected from July 9, 2021, to January 1, 2023. For the pre-vaccine serum, baseline serum was collected from 10 individuals before vaccination with no history of SARS-CoV-2 infection and seronegative for anti-S IgG. The samples were collected as per protocol SDP-003, Human Biological Specimens Collection, dated September 22, 2017, and the qualifications of the principal investigator (Robert Pyrtle, M.D.) were reviewed and approved by the Diagnostics Investigational Review Board (Cummaquid, MA, USA, IRB No.: 202001041A3C).

**Table I.** Characteristics of the sample donors for each group. Age and sample collection after dosage (days) as the mean with the minimum and maximum.

|  | **AstraZeneca**<br>**ChAdOx1nCoV-19** | **Medigen**<br>**MVC-COV1901** | **BNT**<br>**BNT162b2** | **Moderna**<br>**m1273** |
| --- | --- | --- | --- | --- |
| **Vaccine platform** | Adenovector | Protein subunit | mRNA | mRNA |
| **Number of samples** |  |  |  |  |
| **First dose** | 22 | 19 | 18 | 15 |
| **Second dose** | 26 | 22 | 24 | 17 |
| **Third dose** | 23 (heterologous) | 20 (16 heterologous) | 17 | 18 |
| **Sex (% female)** | 72 | 64.7 | 86.7 | 44 |
| **Age (years)** | 36.9 (21–62) | 40.3 (22–61) | 33.4 (21–56) | 47.1 (25–73) |
| **Sample collection after dose (days)** |  |  |  |  |
| **First dose** | 38.5 (25–59) | 26.9 (22–38) | 26.5 (21–39) | 32.9 (22–67) |
| **Second dose** | 26.5 (16–53) | 21.1 (14–27) | 27.3 (18–40) | 36.7 (14–60) |
| **Third dose** | 25 (14–90) | 145 (18–316) | 116.9 (20–325) | 69 (14–260) |
|  |  | 75.7 (17–296) (heterologous) |  |  |
| **Infected individuals (only received third dose)** | 4/23 (17.4%) | 9/20 (45%)<br>7/16 (43.8% heterologous) | 6/17 (35.3%) | 5/18 (27.8%) |

### IgG neutralization titer determination

For each serum sample, the neutralization titer (NT) titer was estimated using the MeDiPro SARS-CoV-2 antibody enzyme-linked immunosorbent assay (Formosa Biomedical Technology Group)according to the manufacturer’s instructions and calculated as previously described^2^ L.

### IgG subclass enzyme-linked immunosorbent assay

The S protein was immobilized at a concentration of 2 mg/mL per well using phosphate-buffered saline (PBS) conjugate on a 96 well polystyrene microplate (R&D Cat No.: # DY990) overnight to estimate the composition of S protein S1-binding IgG subclasses in the serum. A blocking solution comprising 5% bovine serum albumin (BSA) in PBST was added. All samples were diluted 1:10 with 2.5% BSA in PBST under the same conditions. The following mouse anti-human IgG secondary antibodies were used: IgG1 (1:2000; SouthernBiotech, Birmingham, AL, USA), IgG2 (1:500; Southern Biotech), IgG3 (1:500; SouthernBiotech), and IgG4 (1:500; Southern Biotech). The baseline for each subclass was determined by the mean signal of the pre-vaccine sera of seronegative individuals to determine the fold change for the IgG subclass.

### Anti-S IgG purification

BcMag^TM^ Tosyl Activated Magnetic Beads (BioClone, San Diego, CA, USA) were used to purify antigen-specific IgG (S protein) from the serum. For coupling of the S protein, 100 μg of S1 protein (Sino Biological, Beijing, China) was bound to 30 mg of Tosyl magnetic beads. The beads were then divided equally for each sample, with coupling beads containing 2.5 μg of S1 protein used for the serum. Antibodies were eluted with 0.1% trifluoroacetic acid. The S protein-specific IgGs were subsequently analyzed using mass spectrometry.

### Analysis of IgG-Fc glycosylation using mass spectrometry

Tris-HCl (100 mM) was added to 5 µg of each anti-S IgG sample. The samples were reduced with 5 mM TCEP (Sigma-Aldrich, St. Louis, MO, USA) for 30 min at 60 °C and alkylated with 10 mM iodoacetamide (Sigma-Aldrich) in the dark for 35 min at 37 °C. The samples were then digested with 0.2 mg/mL trypsin in 0.1% FA/10% ACN for 16 h at 37 °C, acidified with 0.5% FA and 0.2% trifluoroacetic acid, and desalted with C18 dissolved in 50 µL (50% ACN/DDW). Five milliliters of the solution were inserted into the Speed Vac and dissolved using 6 mL FA (0.1% FA). The tryptic peptide samples were reconstituted in high-performance liquid chromatography (HPLC) buffer A (0.1% formic acid) and loaded across a trap column (Zorbax 300SB-C18, 0.3 × 5 mm; Agilent Technologies, Wilmington, DE, USA) at a flow rate of 15 μL/min in HPLC buffer A. The peptides were eluted using a linear gradient of HPLC buffer B (100% acetonitrile/0.1% formic acid) at a flow rate of 0.18 μL/min and separated on an analytical column (nanoACQUITY UPLC C18 1.7 μm, 0.15 x 10 mm, Waters, Milford, MA, USA) coupled with in-line dilution at a flow rate of 0.1 μL/min HPLC buffer C (0.1% formic acid) for refocusing peptides. Eluted peptides were analyzed with a two-dimensional linear ion trap mass spectrometer LTQ-Orbitrap, controlled by Xcalibur 2.2 software (Thermo Fisher Scientific, San Jose, CA, USA). Intact peptides were detected using full-scan mass spectrometry (MS), performed in the Orbitrap over 400–2,000 Da, and had a resolution of 60,000 at 400 m/z. Accurate m/z values were initially entered as input into the inclusion list, and MS/MS analysis was performed on the LTQ– Orbitrap MS with a 10-ppm tolerance on the parent mass to verify that the given peptides could be detected. 21MS/MS scan events (21 HCD) of 21 target precursor ions for IgG1 with the most abundance were followed by one preview MS scan (Supplementary Table I). The m/z values selected for MS/MS were dynamically excluded for 15 s with a relative mass window of 15 ppm. MS and MS/MS automatic gain control was 1,000 ms (full scan), 300 ms (HCD) or 3 × 10^6^ ions (full scan), and 3 × 10^4^ ions (HCD) for maximum accumulated time or ions, respectively.

### Processing of liquid chromatography with tandem MS data

The signals of the peptides were obtained by analyzing the RAW files with Maxquant 2.0.3.1 software (Max-Planck-Institute of Biochemistry). The intensity of the signal from the precursor was used as the signal. The RAW files were converted into mgf files. The precursor ions were initially selected through manual annotation and literature reports based on mass, charge, and retention time, which was then automated with a custom-written Python script. Python also used mass, charge, retention time, and a scoring system for the fragments of the precursor ion to determine if the precursor contain a glycan. The scoring system was based on the GlyxtoolMS software (12). Fucosylation, galactosylation, and sialylation were estimated by normalizing the sum of the signals of all IgG1 N-297 glycopeptides, as done in previous studies^13,14^ L.

### Measurement of ADCP and ACDC

The assay for ADCP and ACDC were adapted from previous studies^7,15^. Beads (5 × 10^8^) were mixed with 100 ng of the S1-biotinylated subunit (ACROBiosystems; Newark, DE, USA; his-avi tag cat. S1N-C82E8) in FACS buffer (0.5% BSA, 5 nM sodium azide) overnight. Serum plasma (5 µL) was diluted to 1:10 in FACS buffer, and endogenous plasma was inactivated by incubation at 56 °C for 30 min. The inactivated serum or plasma was then incubated with the beads at 37 °C for 2 h to bind antigenic antibodies to the beads. For the ADCP assay, the beads were washed twice with medium for THP-1 cells and then incubated with 1 × 10^5^ cells for 15 h. The cells were then washed using centrifugation at 400 × *g* for 3 min with PBS (2×) and incubated with trypsin EDTA (0.25%; Gibco, Waltham, MA, USA) for 10 min. After two washing steps, the cells were resuspended in 600 µL of FACS and subjected to flow cytometry. For complement activation, the antibody complexes were incubated in 1/50 guinea pig complement (MP Biomedicals, Santa Ana, CA, USA; cat# 086428-CF) in RPMI 1640 medium for 15 min, followed by two washing steps with 15 mM EDTA in PBS. The cells were then incubated with an anti-goat C3 antibody (MP Biomedicals; cat# 0855367) for 15 min, followed by two washing steps and incubation with an anti-goat fluorescent antibody (donkey anti-goat IgG H&L, AF647 Abcam cat# ab150135) for 15 min. The beads were resuspended in 1 mL of FACS buffer for flow cytometry. To measure ADCP, the phagoscore^7^ was calculated as a fraction of bead-positive cells multiplied by the mean fluorescence intensity (MFI, Supplementary Figure 1 A, B). For ADCD, the MFI was used (Supplementary Figure 1 C,D). For both, a fold change was calculated as the ratio in comparison to the phagoscore or MFI to control beads not treated with serum or plasma.

### Statistical analyses

Statistical analysis was performed with R software (ver. 4.3.1) using the dplyr package. The Mann– Whitney U test was used for comparison between two samples. The pairwise Wilcoxon test was used to compare multiple groups. P values < 0.05 were considered significant. The cor.test function in R was used to compare the significance of correlations of the IgG profile parameter with other variables (age, ADCP, ADCD and time interval from injection).The estimated P-value was Bonferroni corrected for the number of comparisons.

## Results

### Study cohort

A total of 272 serum and plasma samples (167 serum, 105 plasma) were collected from 157 individuals vaccinated with the adenovector vaccine ChAdOx1nCoV-19 AstraZeneca (AZ), the protein subunit vaccine Medigen MVC-COV1901, or two mRNA vaccines: m1273 Moderna and BNT162b2 BNT (BioNTech Pfizer) (**Table I**). The samples were collected after administering the first, second, and third doses. For AZ-vaccinated individuals, only samples for the first and second doses were available. For the AZ and Medigen vaccines, heterologous samples were included, with individuals receiving two doses of AZ or Medigen and a third dose of an mRNA vaccine. Most (66.2%) of the samples were obtained from female donors. The age range was 21–73 years.

### The anti-S IgG subclasses change significantly over time in each vaccine group

The NT increased significantly in all vaccines with consecutive doses (**Figure 1A, B**). In Medigen-, Moderna-, and BNT-vaccinated individuals the NT titer increased after the second dose, and AZ-vaccinated individuals after the third dose with a mRNA vaccine. Individuals vaccinated with AZ did not have significant changes in the anti-S IgG subclass level. Anti-S IgG1 levels increased in Medigen-, BNT-, and Moderna-vaccinated individuals after the second dose (**Figure 1A, C**). Anti-S IgG2 also increased significantly in BNT- and Moderna-vaccinated individuals after the second dose and in Medigen-vaccinated individuals after the third dose (**Figure 1A, D**). Anti-S IgG3 did not significantly change in any of the cohorts (**Figure 1A, E**). Anti-S IgG4 increased after the third dose over a prolonged time in mRNA-vaccinated individuals (**Figure 1A, F**). Vaccination with an mRNA vaccine after primary immunization with the AZ vaccine led to a significantly weaker increase in anti-S IgG4 than only mRNA vaccinated individuals. However, heterologous vaccination with two doses of the Medigen vaccine followed by a third dose of an mRNA vaccine led to a strong increase in anti-S IgG4 similar to an mRNA vaccine, in comparison to individuals who received three doses of the Medigen vaccine.

**Figure 1:**
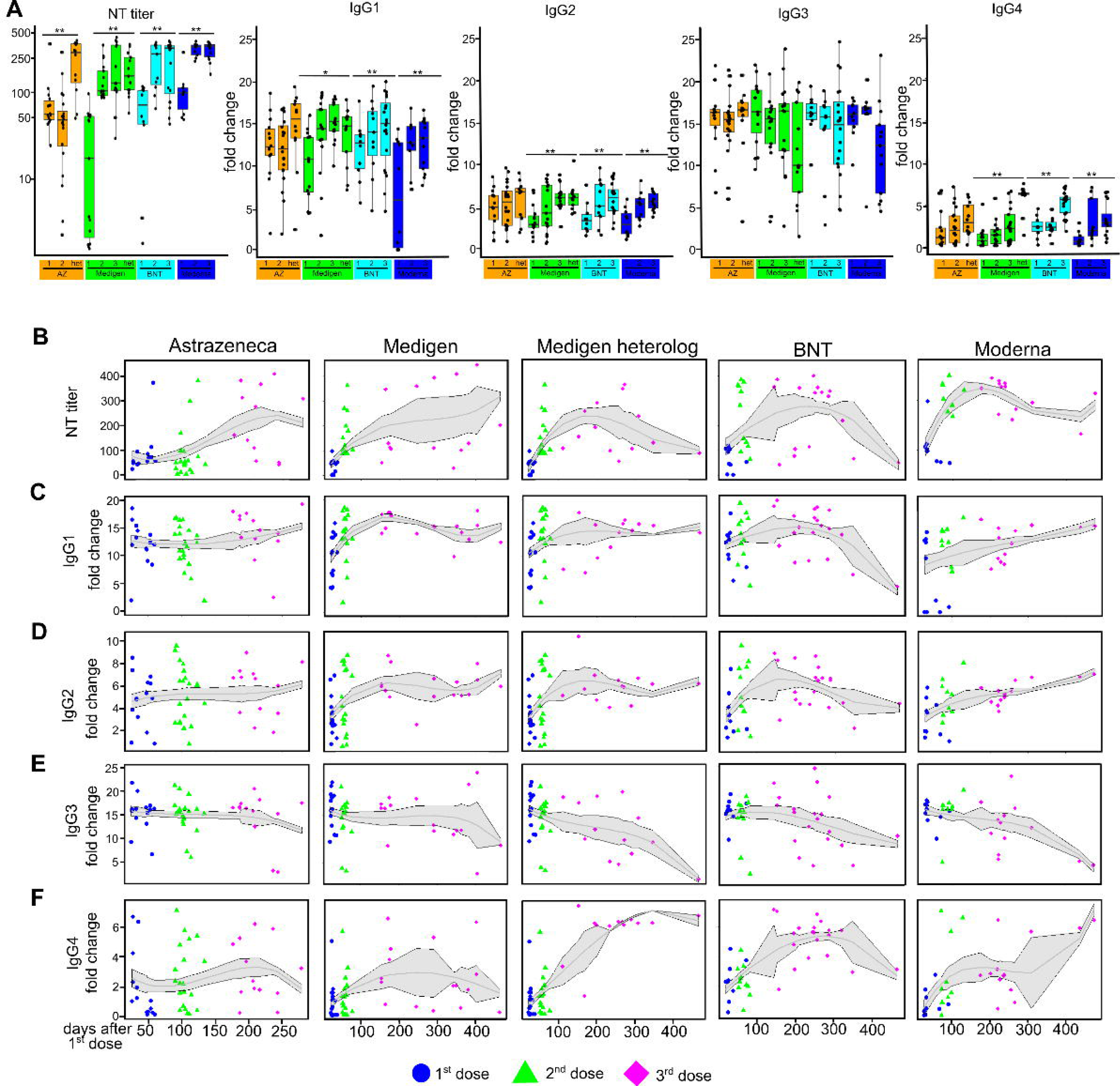
Neutralization titer (NT) and anti-spike (S) IgG1, 2, 3, and 4 subclass levels by doses with significance level * P<0.05,** P<0.01 (Wilcoxon–Mann–Whitney test) **(A)** and from time after application by dose (**B–F**). The anti-S IgG subclass level was defined by the fold change compared to the mean anti-S IgG subclass level of 10 non-vaccinated seronegative individuals. het: heterologous (first to second dose non-mRNA vaccine, third dose mRNA vaccine)

### IgG Fc-glycosylation is stable over time

n-297 glycosylation of anti-S IgG was measured using liquid chromatography with tandem MS. The mean levels of fucosylation, galactosylation, bisection, and sialylation were 97.8, 68.5, 5.3, and 8.4%, respectively (**Figure 2**). No significant differences were observed between the different vaccine types apart from BNT, which had a significantly higher galactosylation than Medigen (**Figure 2A, C**). The glycosylation level was stable over time.

**Figure 2:**
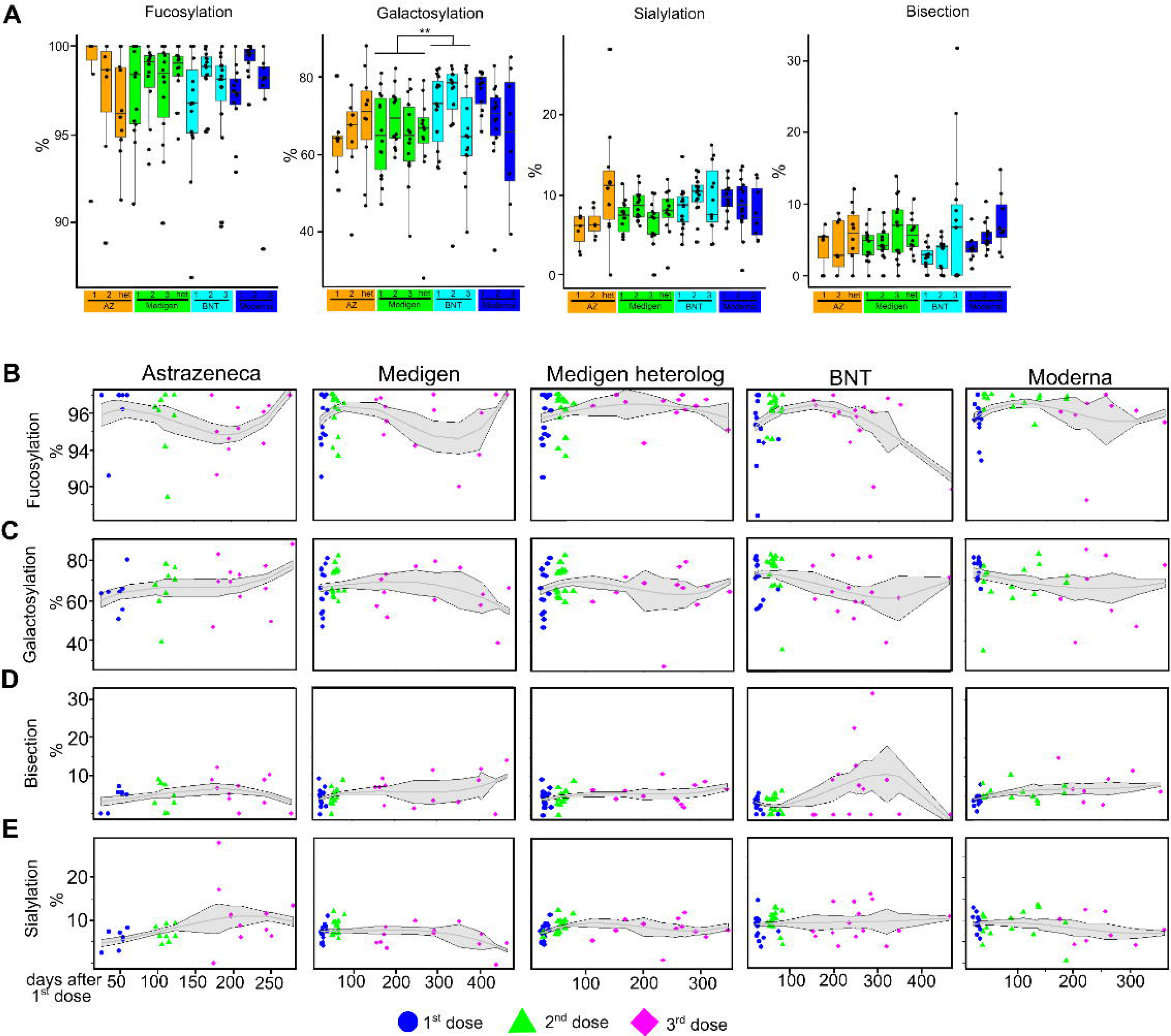
Anti-S IgG n-297 Fc-glycosylation for fucosylation, galactosylation, bisection, and sialylation by doses with significance level, *P<0.05, **P<0.01 **(A)** and from time after application by dose (**B–E**).

### The IgG profile differs significantly from the first to the third dose in all vaccine groups, except in AZ-vaccinated individuals

The IgG subclass and NT titer correlated significantly with each other, except for IgG3 (**Figure 3A**). The NT titer was further correlated with bisection and anti-S IgG sialylation. Anti-S IgG galactosylation correlated with anti-S IgG sialylation. Principal component analysis identified that galactosylation, sialylation, and IgG subclasses 1, 2, and 4 represent the main variables explaining the variance (**Figure 3B**). In each vaccine cohort, the third dose is significant in its IgG profile from the first dose, except for in AZ vaccinees (**Figure 3C–F**). In Medigen vaccinated each dose was significant from each other.

**Figure 3:**
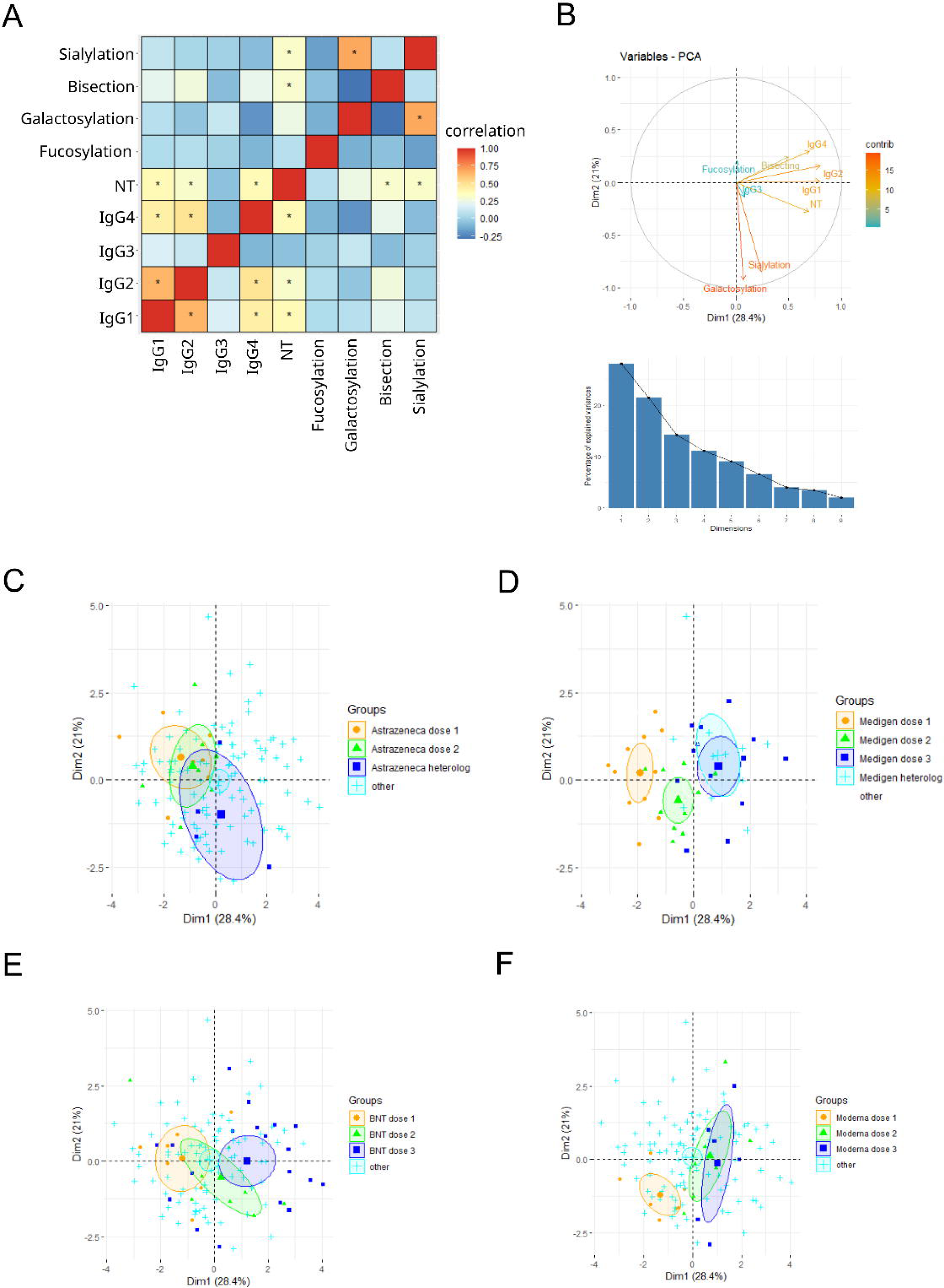
Pearson’s correlation of the parameters of the anti-S IgG subclass level and the anti-S IgG Fc-glycosylation across all samples. A significant correlation between the IgG 1, 2, and 4 subclass level and galactosylation and sialylation was observed, *P<0.05 (**A**). Principal component analysis of all samples among anti-S IgG subclasses and glycosylation with the contribution of the variables for the first and second dimension (**B**). The different vaccinated individuals are divided by vaccine and dose for the first two principal components. The circles present a 95% confidence interval (**C–F**).

### Specific sex differences occur only in BNT-vaccinated individuals after the third dose

After the third dose of the BNT vaccine, all female individuals had higher anti-S IgG1, 2, and 4 titers than males (**Figure 4A–C, E, Supplemental Figure 2A–C, E**). Furthermore, anti-S IgG Fc-galactosylation decreased after the third dose, whereas bisection increased in females but not in male BNT vaccinated individuals (**Figure 4G, H; Supplemental Figure 2G, H**). Bisection was significantly higher in females after each dose than in male BNT-vaccinated individuals. However, no sex-based differences were observed in individuals vaccinated with other vaccines (**Figure 4, Supplemental Figure 2**).

**Figure 4:**
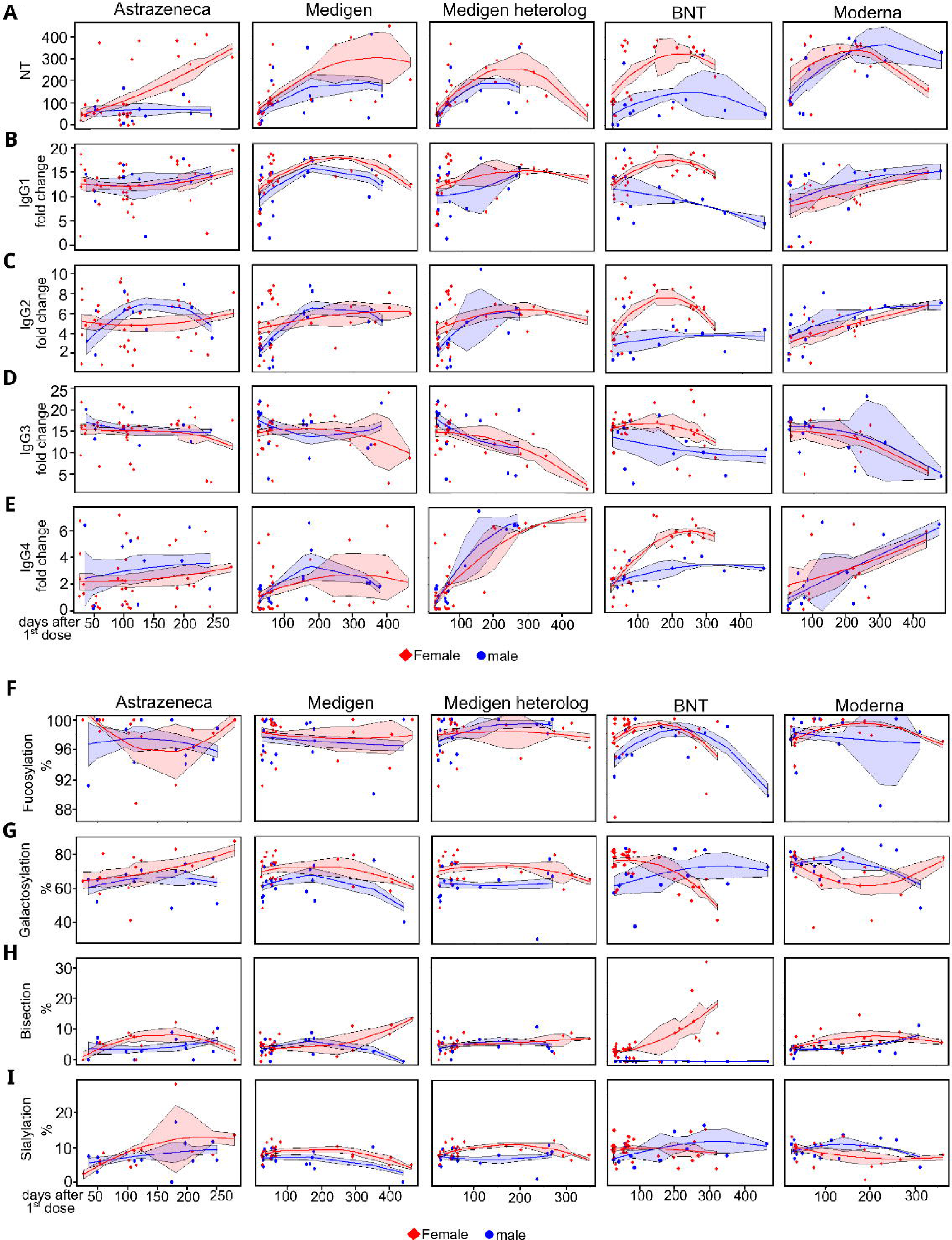
NT titer, anti-S IgG subclass level (**A–E**), and anti-S IgG glycosylation (**F–I**) separated by sex and days after the first dose. In BNT-vaccinated individuals, females have significantly higher NT titers and IgG1, 2, and 4 than males. Galactosylation and bisection are significantly different in BNT-vaccinated individuals than in male BNT vaccinated groups. Significant levels by dose are presented in **Supplemental Figure 2**.

### ADCP declines in all vaccine groups over time, and ACDC is elevated specifically in protein subunit-vaccinated individuals

ADCP and ADCD were measured by fluorescence bead-based assays and flow cytometry (**Supplementary Figure 1**). ADCP and ADCD were not significantly correlated to any IgG subclass titer or Fc-glycovariants (**Figure 5A**). No significant difference was observed in the ADCP capacity of the serum against S-protein immunocomplexes between the different vaccinated groups (**Figure 5B, D**). However, ADCP significantly declined in all vaccines after the third dose. In BNT-vaccinated individuals, ADCP declined after the second dose. ADCD increased in Medigen-vaccinated individuals after three doses, showing a significant increase in ADCD capacity (**Figure 5C, E**). AZ-vaccinated individuals had a significantly lower ADCD compared to the other vaccinated groups and were not significantly different from baseline complement activity (**Figure 5C**).

**Figure 5:**
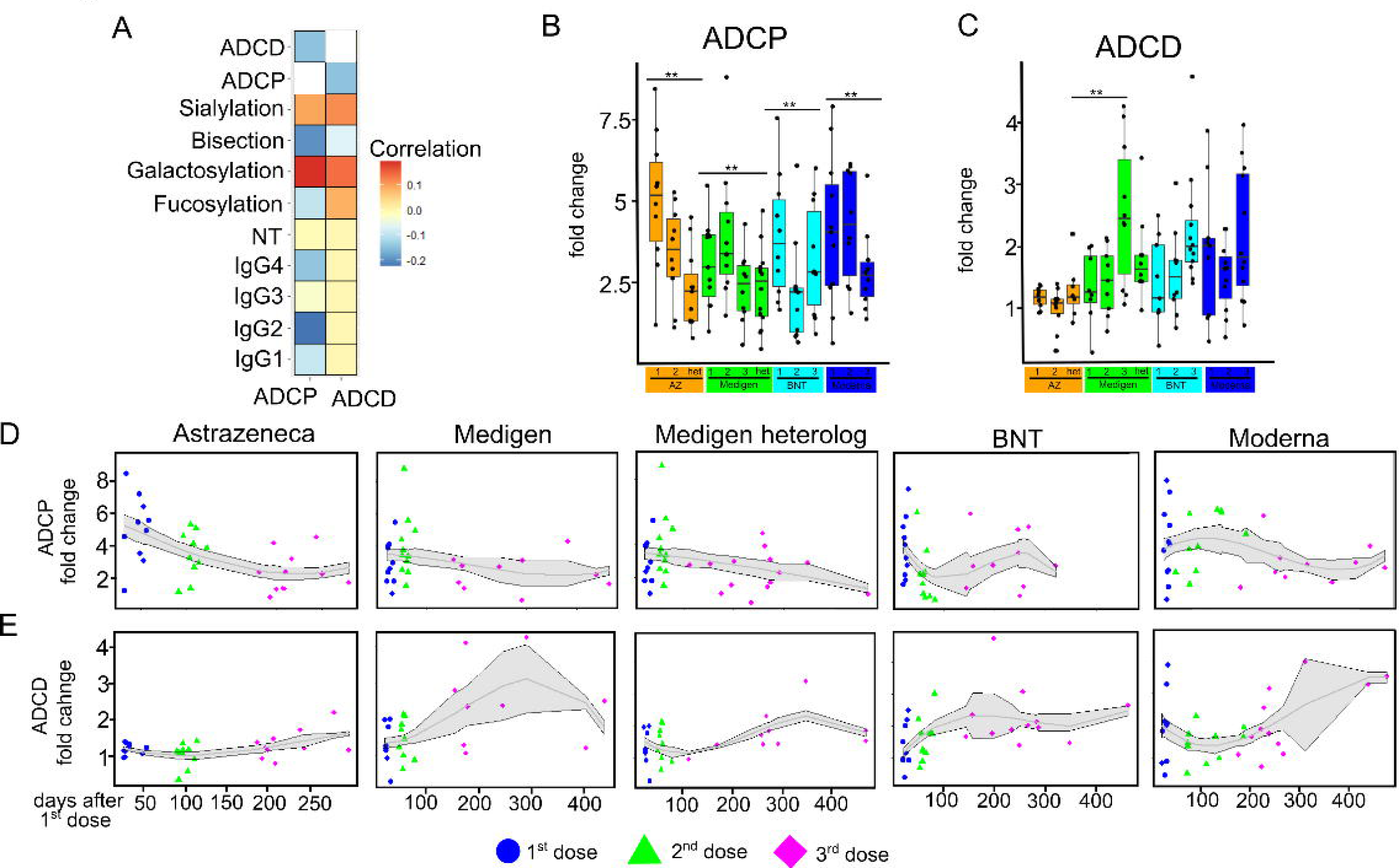
Antibody-dependent phagocytosis (ADCP) (**A, D**) and antibody-dependent complement deposition (ADCD) (**B, E**) as a fold change compared to samples treated without serum or plasma from vaccinated individuals. Correlation of ADCP and ADCD to anti-S IgG subclass and Fc-glycosylation (**C**).

### Influence of COVID-19 infection, age, and time after each dose

No major COVID-19 outbreaks occurred in Taiwan before January 2022. Therefore, the sample collection only included COVID-19-recovered patients after the third vaccination dose. No significant differences were observed in the vaccinated cohorts between recovered COVID-19 patients and non-infected vaccinated individuals (**Supplemental Figure 3**). However, the number of infected individuals for each subgroup was low (only one or two patients in some subgroups). Therefore, the dataset was too small to determine the role of SARS-CoV-2 infection after the third vaccination dose. Furthermore, none of the IgG structural features or antibody functions significantly correlated with age except for anti-S IgG Fc-fucosylation, which was significantly negatively correlated with age in individuals vaccinated with Moderna after the second dose (**Supplemental Figure 4A**). In terms of the time interval after the sample was taken after each dose, anti-S IgG-galactosylation had a significant negative correlation in BNT-vaccinated individuals after the second dose, and anti-S IgG2 was significantly positively correlated after the third dose in Moderna-vaccinated individuals (**Supplemental Figure 4B**).

## Discussion

This study is the first to investigate the IgG profile and antibody function of a protein subunit COVID-19 vaccine with long-term data and the effect of heterologous combinations with mRNA vaccines. Each vaccine type showed unique long-term changes after three doses.

Viruses based on natural infection or adenovector vaccines as primary immunization event can prevent the increase of anti-S IgG4 after mRNA vaccination. In agreement with previous studies, an increase in anti-S IgG4 after the third vaccination dose of mRNA was observed. Primary immunization with the AZ vaccine impeded the increase in anti-IgG4. In contrast, primary vaccination with two Medigen doses and a third dose of mRNA vaccines increased anti-S IgG4 after the third mRNA vaccine dose^7–9^. The non-enveloped replication-deficient DNA virus used in AZ and the enveloped SARS-CoV-2 RNA virus have different modes of infection; however, they exhibited a similar effect in primary immunizations in later mRNA vaccines. The main adjuvant of the Medigen vaccine is CPG aluminum-hydroxide, which activates the innate immune receptor TLR9 for DNA. This receptor is also activated by the DNA adenovector^16,17^. However, adenovectors can also activate the immune system through TLR9-independent mechanisms^17^. Therefore, more research is needed to determine which combination of factors leads to the different outcomes in the long-term effects of the IgG subclasses. A study on mice showed that the germinal center of the IgG subclass scarcely changes after primary immunizations^18,19^. This finding indicates that the first immunizations determine the development of further exposures to the antigens. Thus, since all vaccines are based on the full-length S protein, the distinct features of their IgG must be due to other factors, most likely the immunostimulatory effect of other components and the biodistribution of the vaccines.

ADCP activity declined in all vaccines after the third dose, which is consistent with another study on BNT^20^. This finding is correlated with the anti-S IgG2, and IgG4 increases because both have low affinities for Fc receptors and lower Fc-mediated functions like ADCP. A negative correlation was observed with the anti-S IgG2 and 4 levels; however, this relationship was not significant. This finding is in contrast to another study, in which the decrease in ADCP activity was associated with an increased anti-S IgG4 titer^7^, possibly due to different normalization methods. The previous study normalized the samples to the IgG concentration, whereas the samples were normalized to the plasma volume in this study, which is more commonly used^20–22^ and is representative of the overall humoral immune response. Furthermore, normalization of the antigenic IgG concentration is more suitable for antibody therapies in which the applied antibody concentration can be controlled; however, ADCP decline may indicate a weakening of antibody-mediated protection since ADCP and ADCC are potential protective mechanisms of Fc-mediated functions against viruses^23^.

In contrast to another study, ADCD did not significantly change in individuals vaccinated with the mRNA vaccines^7^. A novel finding of this study was the increase of ADCD after three doses of Medigen, but not if the individual was also vaccinated with two doses of a protein vaccine and a third mRNA vaccine. However, the mechanisms driving this difference are unknown. One potential reason is the CPG adjuvant that alters the complement-dependent response^24^. The complement system has no potential protective role in viral or SARS-CoV-2 infection, and overactivation of the complement system may be harmful^23^. No adverse reactions were reported in the three-dose Medigen-vaccinated population if these individuals were infected with SARS-CoV-2.

This study showed distinct features between the Moderna and BioNTech/Pfizer mRNA vaccines. BNT was the only vaccine with a long-term difference between female and male vaccinated individuals. This finding is in accordance with other studies, which also found a higher anti-S titer in females after the third BNT dose^25,26^. Because both vaccines are based on mRNA and encode the same protein, and Moderna had a higher dose of mRNA of 100 µg compared to 30 µg in BNT, other factors may be involved in this difference. One major factor that must be investigated is the effect of the lipid nanoparticles since they both comprise a significantly different set of lipid nanoparticle (LNP) components^27^. Some LNP formulations have a slower clearance in women than in men, and phagocytosis and macrophage activation are stronger in females than in males^28^ (28). Furthermore, an LNP-mRNA formulation showed a stronger inflammatory reaction in female rabbits than in males^29^. However, no study has investigated the influence of LNPs on the long-term differences in IgG glycosylation in individuals vaccinated with mRNA vaccines. There is only one study which found Sialyltransferase 1 is expressed the higher in B-cells after immunizations with oil-based compared to other adjuvants^30^.

This study has some limitations. The samples were not collected from the same individuals for each dose. Therefore, precise individual based kinetics of the IgG profile cannot be made. Since it was an observational study, the groups are not controlled for demographic characteristics, interval of sample collection to the last vaccine dose and infection status and the sample size was too small to assess the influence of demographic characteristics beyond sex-based differences.

In conclusion, this study showed specific long-term changes in the IgG profile for three COVID-19 vaccines after three doses and heterologous vaccination with protein subunit and mRNA vaccines. Comparing these different vaccine platforms enables the selection of future immunization schemes to achieve the desired adaptive immune response successfully.

## Data Availability

The samples were collected as per protocol SDP-003, Human Biological Specimens Collection, data September 22, 2017 and qualifications of the principal investigator (Robert Pyrtle, M.D.) were reviewed and approved by the Di- agnostics Investigational Review Board (Cummaquid, MA, USA).

## Acknowledgments

We thank the Clinical Proteomics Core laboratory of the Chang Gung Memorial Hospital, Linkou for performing the LC-MS/MS of our prepared samples. This work has been supported by NIH NIAID U01AI151698, United World Antiviral Research Network, part of the NIAID CREID network. This work was also supported in part by the Research Center for Emerging Viral Infections from The Featured Areas Research Center Program within the framework of the Higher Education Sprout Project by the Ministry of Education (MOE) in Taiwan. Additionally, we received support from the Ministry of Science and Technology (MOST), Taiwan (grant number: MOST 111-2634-F-182-001, MOST 109-2327-B-182-002), and the Chang Gung Memorial Hospital (grant number: CORPD1K0011-12).

