## Supplemental Figures and Table for "Specific long-term changes in anti-SARS-CoV-2 IgG modifications and antibody functions in mRNA, adenovector, and protein subunit vaccines"

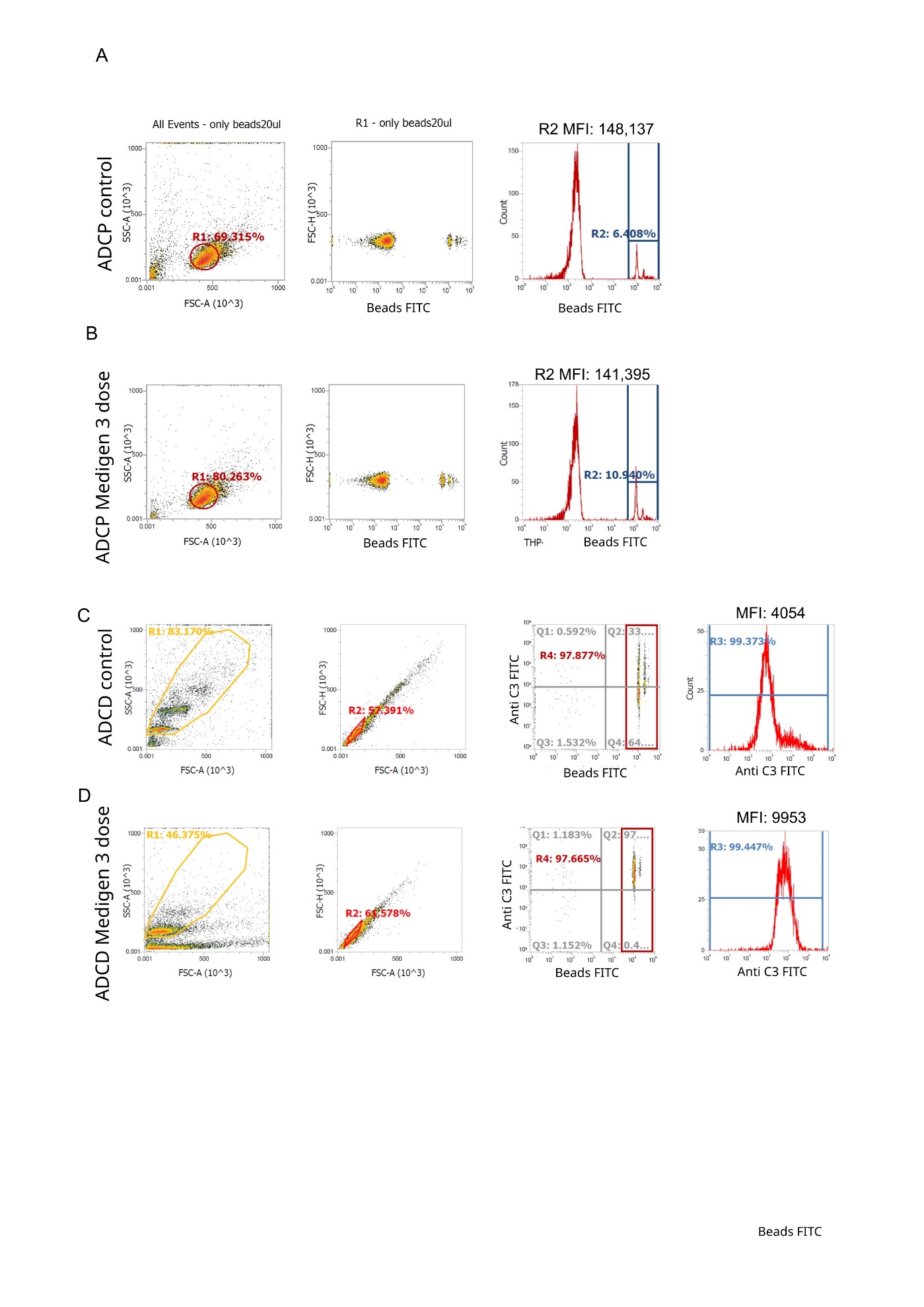

**Supplemental Figure 1**: Flow cytometry of the ADCP (**A,B**) and ADCD assays (**C,D**) from control beads coated with S1 protein without serum or plasma samples from vaccinated individuals (**A,C**) or treated with a plasma sample from a vaccinated individual (**B,D**). In the ADCP assay, THP-1 cells are identified by their size in FSC and SSC scatter. Cells that have phagocytosed beads have a higher fluorescence and can be counted as positive cells. The phagoscore will then be calculated by multiplying the fraction of positive cells with the mean fluorescence intensity (MFI) of the positive cells. The ratio of the phagoscore of the sample to the control will be the fold change. In the ADCD assay, beads are identified by their size and yellow fluorescence. The complement deposition is measured by the MFI of the channel for the anti-C3 staining. The ratio of the MFI between the sample and the control will be the fold change.

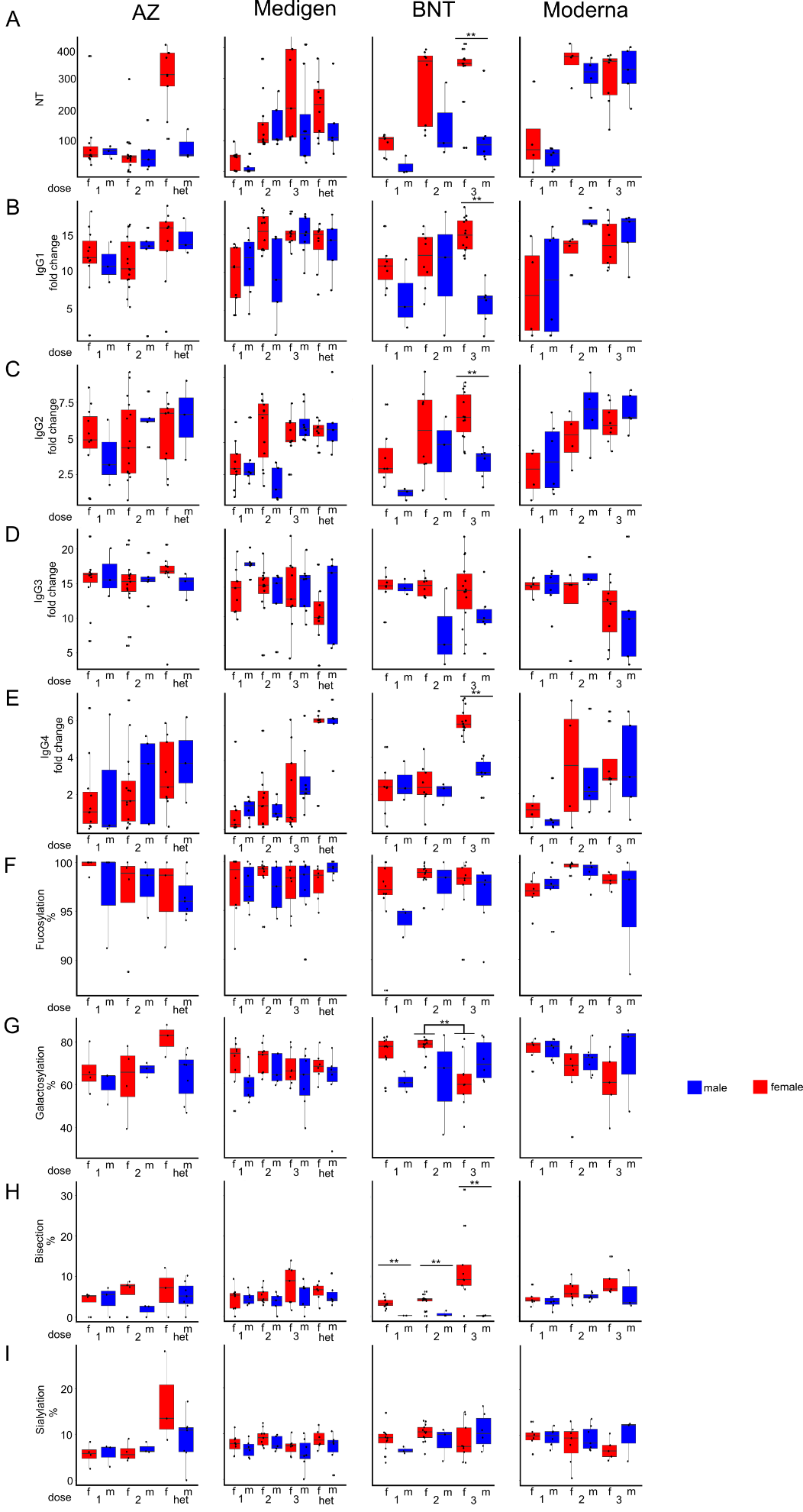

**Supplemental Figure 2**: Sex based differences of anti-S IgG NT titer (**A**) subclass (**B-E**) and anti-S IgG glycosylation (**F-I**) in the different vaccine groups by dose (male =blue, female=red). * p<0.05,** p<0.01

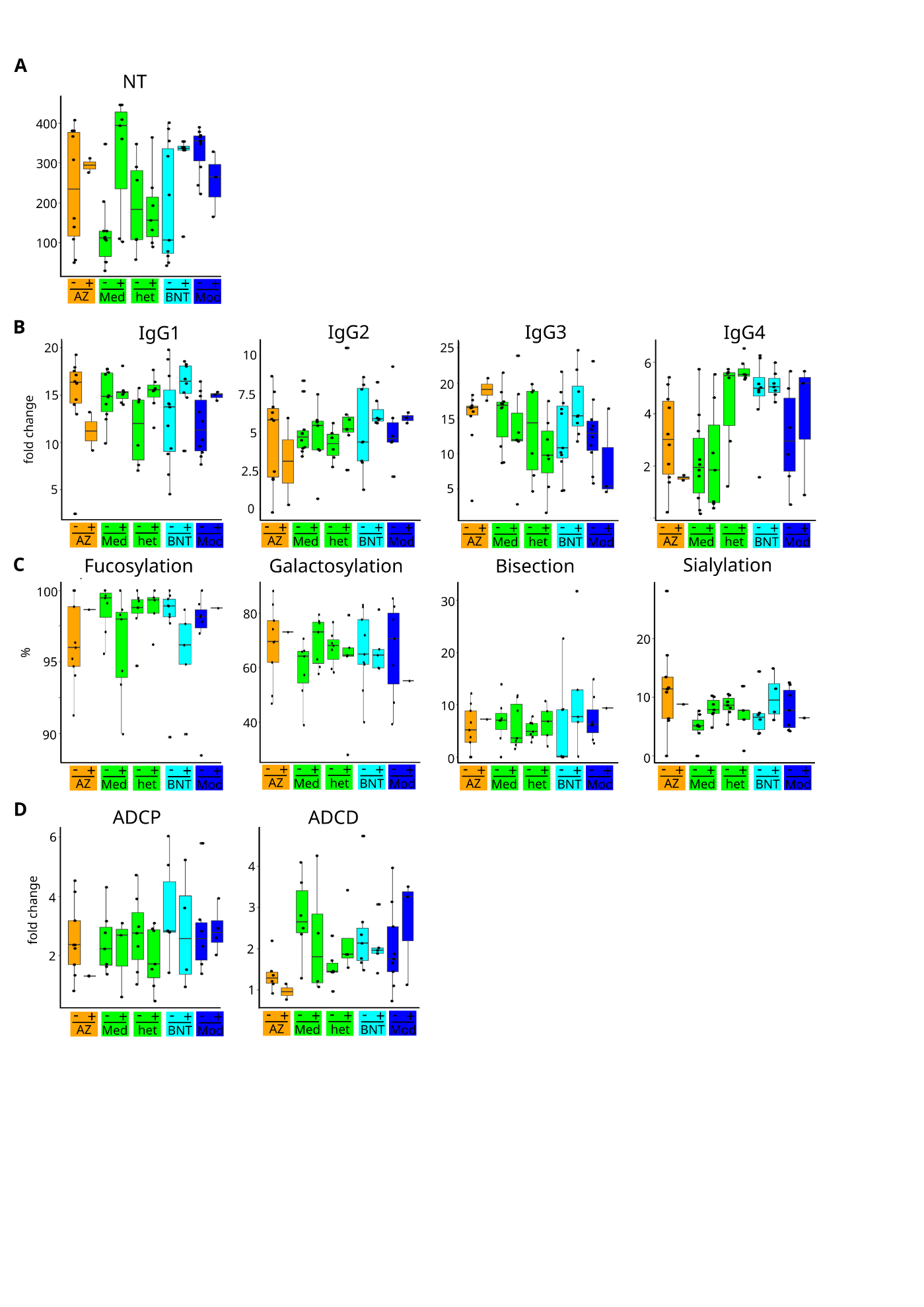

**Supplemental Figure 3**: Comparison of Anti-S NT titer (**A**) , anti-S IgG subclass (**B**), Fc-glycosylation (**C**) and antibody functions (**D**) between COVID-19 recovered and non-infected individuals after the 3^rd^ dose. - : not infected, + : infected, Med: Medigen, Mod: Moderna

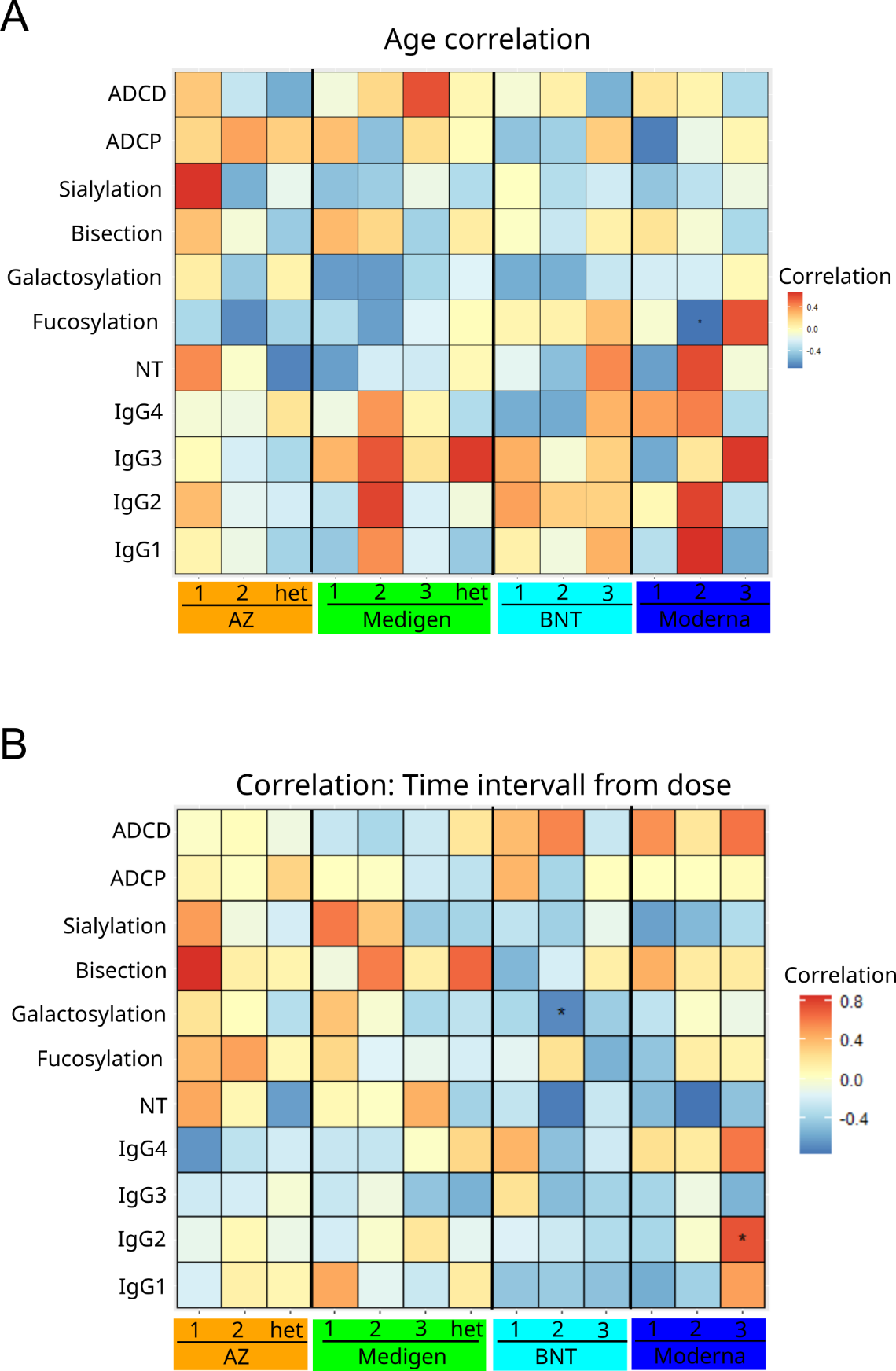

**Supplemental Figure 4**: Correlation of the NT-titer, Anti-S IgG subclass and Fc-glycosylation with for each vaccinated group and dose with age (**A**) and the time intervall after each dose (**B**). * p<0.05

**Supplemental Table 1**: Glycan structure from n-297 glycosylation analyzed by LC-MS/MS mode and their mass/charge (m/z) for for 2+ and 3+ charge. Nomenclature, F: Fucosylated; G0, G1, G2: Galactose residues (0, 1 or 2 Galactose residues). S, S2: Sialylation (1 or 2 sialic acid residue). N: Bisection

Fucose (F)

Galactose (G)

Sialic acid (S)

N-acetylglucosamine

Mannose

| Glycan structure | Abbreviation | Precursor m/z per charge | |
| --- | --- | --- | --- |
|  |  | 2+ | 3+ |
| 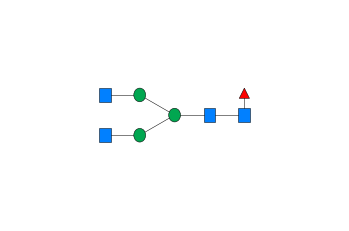 | G0F | 1317.52 | 878.68 |
| 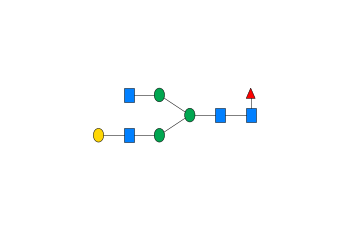 | G1F | 1398.55 | 932.7 |
| 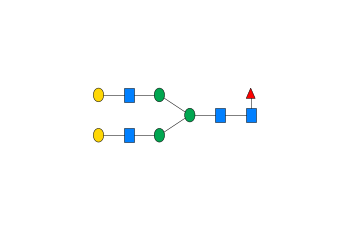 | G2F | 1479.58 | 986.72 |
| 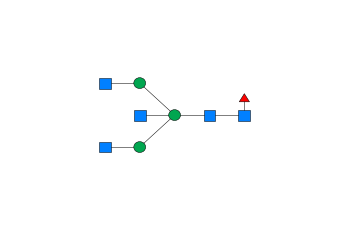 | G0FN | 1419.07 | 946.38 |
| 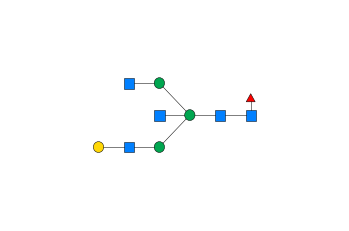 | G1FN | 1500.09 | 1000.4 |
| 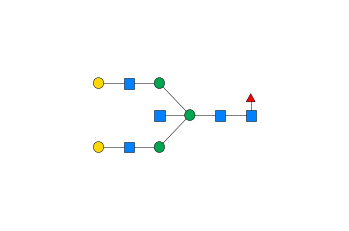 | G2FN | 1581.12 | 1054.42 |
| 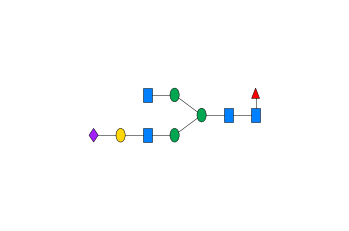 | G1FS | 1544.1 | 1029.74 |
| 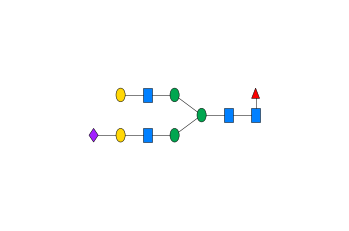 | G2FS | 1625.13 | 1083.75 |
| 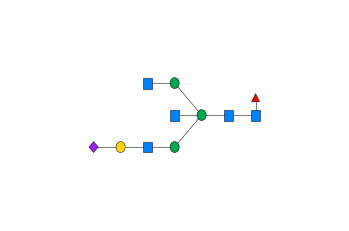 | G1FNS | 1645.64 | 1097.43 |
| 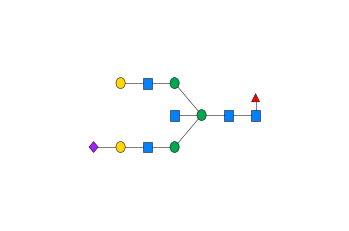 | G2FNS | 1726.67 | 1151.45 |
| 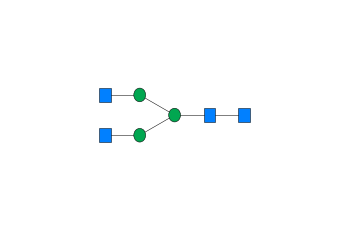 | G0 | 1244.5 | 830.0 |
| 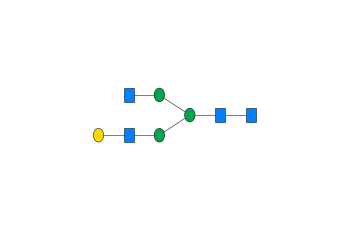 | G1 | 1325.52 | 884.02 |
| 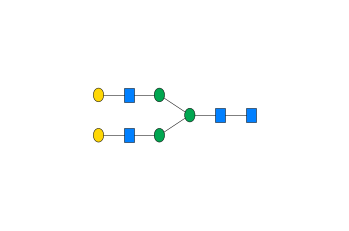 | G2 | 1406.55 | 938.04 |
| 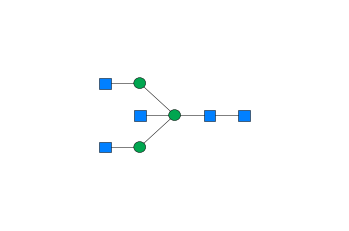 | G0N | 1346.04 | 897.69 |
| 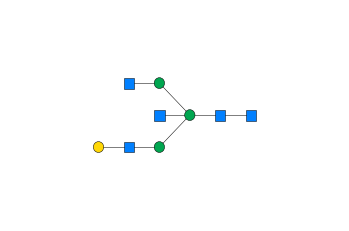 | G1N | 1427.06 | 951.71 |
| 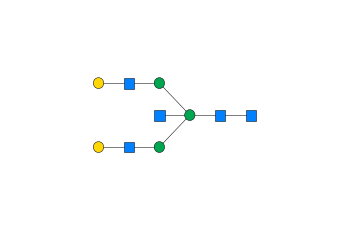 | G2N | 1508.09 | 1005.73 |
| 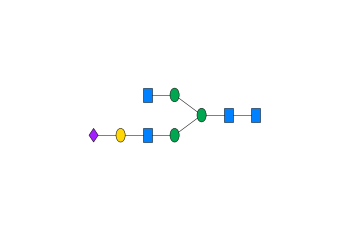 | G1S | 1471.07 | 981.05 |
| 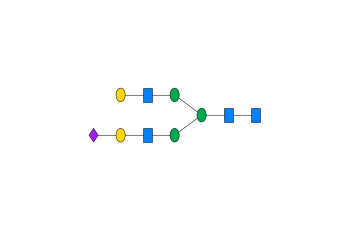 | G2S | 1552.1 | 1035.07 |
| 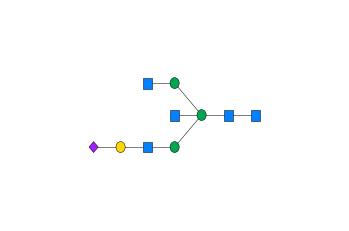 | G1NS | 1572.61 | 1048.74 |
| 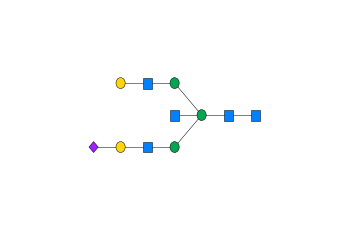 | G2NS | 1653.64 | 1102.76 |
| 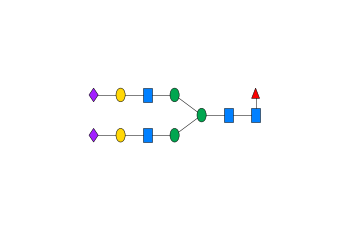 | G2FS2 | 1770.67 | 1180.79 |
